# Have deaths of despair risen during the COVID-19 pandemic? A rapid systematic review

**DOI:** 10.1101/2022.04.05.22272397

**Authors:** Hania Rahimi-Ardabili, Xiaoqi Feng, Phi-Yen Nguyen, Thomas Astell-Burt

## Abstract

**Objective:** To systematically review the literature on the impact of the COVID-19 pandemic on deaths of despair (suicide, overdoses and drug-related liver diseases).

**Methods:** Five electronic databases were searched using search terms on deaths of despair and COVID-19.

**Results:** The review of 70 publications included indicates that there is no change or a decline in the suicide rate during the pandemic compared to the pre-pandemic period. Drug-related deaths such as overdose deaths and liver diseases, however, have been increased compared to the pre-pandemic rate. Findings are mainly from middle-high- and high-income countries and data from low-income countries are lacking. Synthesis of data from subgroup analysis indicates that some groups such as Black people, women and younger age groups would be more vulnerable to socioeconomic disruption during the pandemic.

**Conclusion:** Studies included in this review were preliminary and suffered from methodological limitations such as lack of inferential analysis or using provisional data. Further high-quality studies are needed considering the contribution of factors such as disease prevalence, government intervention and environmental characteristics.

## Introduction

Beyond the acutely devastating rise in communicable disease mortality, impacts of the protracted socioeconomic disruption unleashed by the COVID-19 pandemic on population health are still emerging ^1-5^. Early reports include potential aggravation of depression and anxiety ^1,2^, increases in suicidal ideation and behaviour ^3,4^, and drug overdoses ^5^. These preliminary findings align with epidemiological studies of previous economic downturns, such as the global financial crisis of 2008-9, which had dire consequences for population health and health equity ^6,7^. While some health impacts may be concurrent with crisis (e.g. stress), others manifest over time as biopsychosocial risk factors such as job loss, food insecurity, precarious housing availability, death of a loved one, and exposure to violence accumulate and in some cases overcome individual resilience ^8^, This quantum of social determinants commonly experienced during economic downturn can induce and aggravate a sense of despair (derived from *desperare*, ergo ‘down from hope’ ^9^) that undermines individual and shared meaning-making ^10^. Despair, often in concert with concomitant factors such as loneliness, which is thought to have been aggravated by social isolation practices enacted to perturb the spread of COVID-19, may lead to future discounting of maladaptive behaviours (e.g. alcoholism, substance misuse) and increased risks of death from drug-related poisoning, liver diseases and suicide ^11^. Case and Deaton coined the phrase ‘deaths of despair’ to describe these causes of death, and first reported an increase in deaths of despair for non-Hispanic middle-aged White working class in the US in 2015 ^11,12^. Since then, other studies have indicated similar increases in other ethnic groups and countries ^11-13^. Based on prior evidence an increase in deaths of despair induced by the COVID-19 pandemic is highly plausible. But while some work has reported deaths of despair rising in the US in 2020 above pre-COVID-19 levels ^14^, there remains no systematic review of the literature to determine if this is an isolated case, or whether it is reflective of wider trends. This rapid systematic review aims to resolve that gap in knowledge.

## Materials and Methods

This review was conducted according to the Preferred Reporting Items for Systematic Review and Meta-Analyses (PRISMA) guidelines for systematic reviews ^15^. Study outcomes were defined based on Case and Deaton’s definition of death of despairs, i.e. suicide (ICD10 X60-84, Y87.0), poisonings (ICD10 X40-45, Y10-15, Y45, 47, 49), and alcoholic liver diseases and cirrhosis (ICD10 K70, K73-74). ‘*Poisonings are accidental and intent-undetermined deaths from alcohol poisoning and overdoses of prescription and illegal drugs*’ ^11^.

### Study Selection

Articles were included if they evaluated the deaths of despair during the COVID-19 pandemic. Due to the sensitivity of COVID-19 subject, studies that have been published as editorials and letters to expedite the publication process are included if they have used original objectively collected data. Table 1 outlines detailed inclusion and exclusion criteria.

**Table 1.**
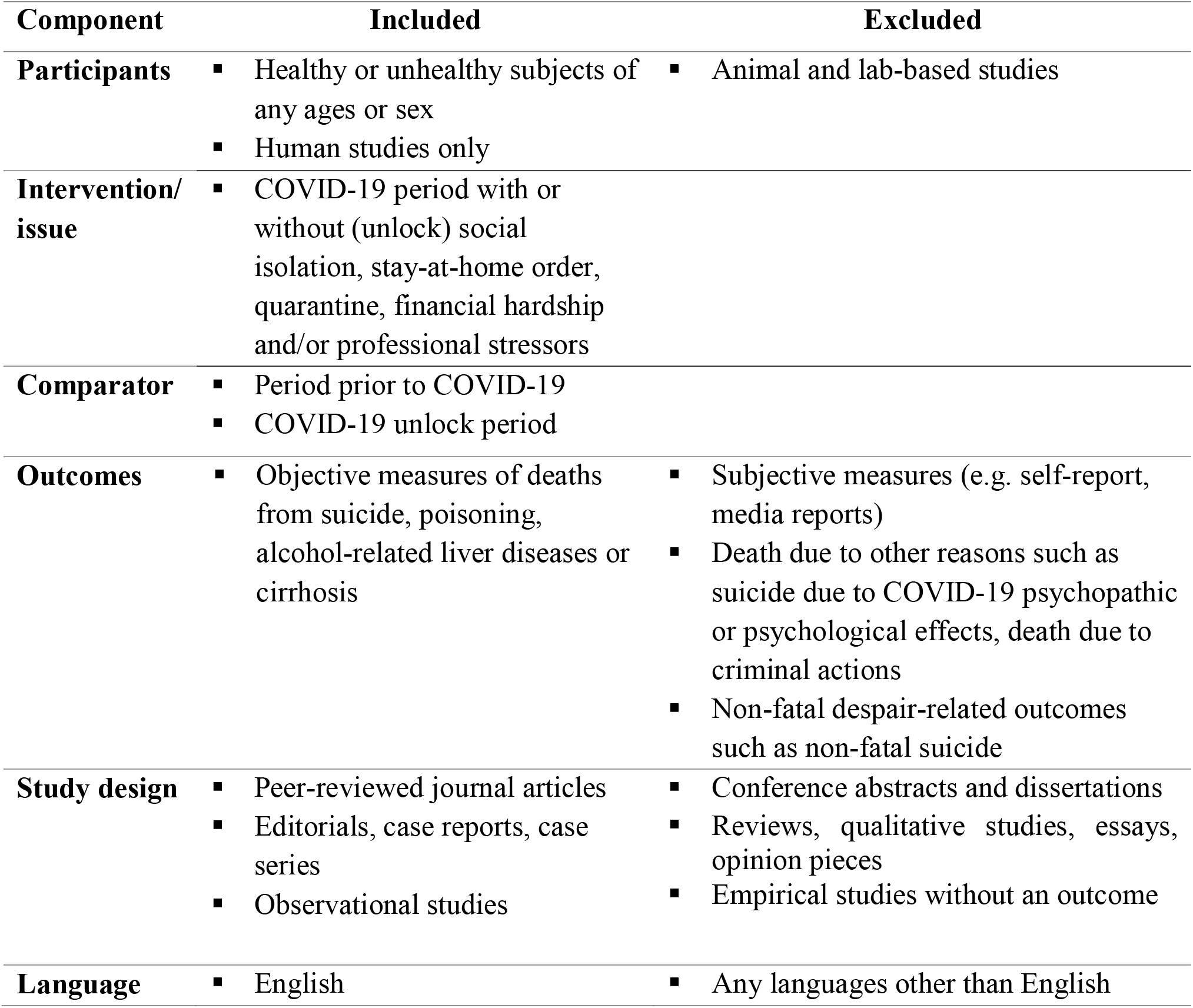
Inclusion and exclusion criteria of the review

### Search Strategies

The following electronic databases were searched on 29 Aug 2021: MEDLINE, EMBASE, Scopus, CINAHL and PsycINFO. Search Strategy partially adapted from previous systematic literature reviews ^3,6^. The COVID-19 search strings were used when there was no related filter available within the database using strings developed by librarians ^16,17^. We used term keyword combinations of ‘despair and ‘deaths’ and ‘COVID-19’ searched in titles and abstracts. See Table S1 for the complete search strategy.

Study selection was completed via a two-step screening process using Covidence software (Veritas Health Innovation, Australia). Two reviewers () independently screened title/abstracts then full-texts to identify eligible articles. Any disagreements were resolved by reviewing full-texts in the title/abstract screening stage and by discussion among investigators in the full-text screening stage. The reference lists of the relevant articles were also reviewed by one reviewer () to identify any other eligible studies that were missed in the initial search process.

### Data Extraction

One author () extracted and synthesised data from the included articles into an Excel sheet. The extracted data included author information, year of publication, study area, study design, population, and sample size. In addition, we collected and synthesised data on the period that the data was collected, methods used to measure outcomes, outcomes (with ICD 10 if reported), comparison period, statistical analysis, covariates adjusted, main results, and mediating and moderating factors if assessed. Comparison time periods are divided into two categories of 1) a period leading to the pandemic (e.g. Jan - Feb 2020 vs Mar – Apr 2020) or 2) the same time period of the previous year (s) (e.g. Mar-May 2019 vs Mar-May 2020). Countries are classified into two categories of low to upper-middle-income and high-income countries based on WHO definition ^18^. The direction of changes in death outcomes between COVID-19 and pre-COVID-19 periods are presented as increased, decreased, or no change. The majority of studies defined their cut-point for pandemic according to the date/month that the state of emergency was declared or lockdown measures introduced.

### Quality Assessment

The National Heart, Lung, and Blood Institute quality assessment tools were used to evaluate the qualities of ecological and cross-sectional l included articles ^19^. Three further items have been added for ecological studies ^20^. For case-report and case series studies, critical appraisal tools developed by Joanna Briggs Institute were used ^21^. For each item in the list, three options for answers are suggested, which are ‘Yes’, ‘No’ or ‘Other’ (NR, NA). If the criteria were met (Yes) it is assigned to the value of 1, otherwise, 0 points were assigned. The scores below 50, between 50 and 74 or above 75 meant the articles were regarded as low, fair and high quality, respectively. The same classification was also used previously ^22^. One reviewer () conducted the quality assessment.

## Results

After removing duplicates, 2,490 articles remained for title/abstract screening. Full texts of 182 articles were reviewed, and 70 articles were selected for this systematic review. Of 70, 3 articles described two different outcomes, and one article ^23^ was an update of an earlier study ^24^. 40 studies were on suicide deaths, 30 on overdose deaths, two on alcohol-related liver disease deaths, one on hanging and poisoning (all intent) deaths. Studies were either funded by public organisations or had no funding. Figure 1 summarises the search process.

**Figure 1.**
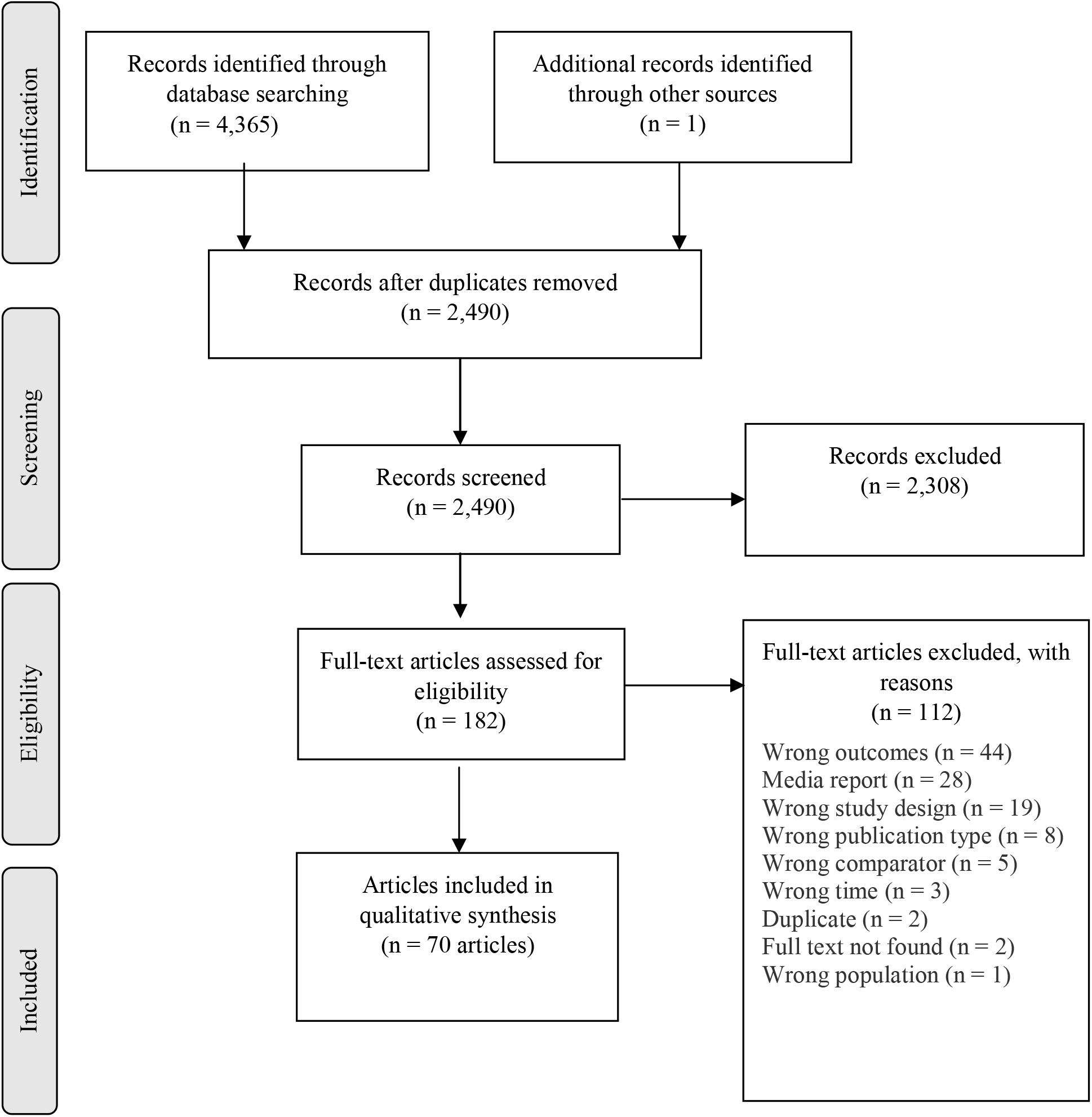
PRISMA flow diagram

### Study characteristics

Most of the articles (80%, n = 56) were published in 2021. Seventeen studies may not have gone through a peer-review process (e.g., editorial) ^23-40^. Almost every study analysed objective data, but one which utilized a survey to collect data about student suicide rates from universities ^31^.

Studies were mainly ecological or cross-sectional in design (n = 61), and a few were case-report/case series studies or studies that only provided descriptive information (n = 9). The COVID-19 study period varied from one month to one year. Overall, 17 countries were included. Some countries were studied several times, such as the US (n = 28) and Japan (n = 11), considering different states, populations, or COVID-19 period. Figure 2 demonstrates the geographical variation of the studies included. Studies were mainly targeted the general population or adults, except for some that examined specific settings such as hospitals or specific populations such as children. There was no consistent pattern in the time compared (i.e period preceding COVID-19 or period at the same time of previous years). The study characteristics and findings are summarized in Tables S2 and S3 for suicide deaths and drug-related deaths, respectively. Over half of the studies (n = 38) were judged to be of low quality, with only eight studies rated as high quality (Table S4)

**Figure 2.**
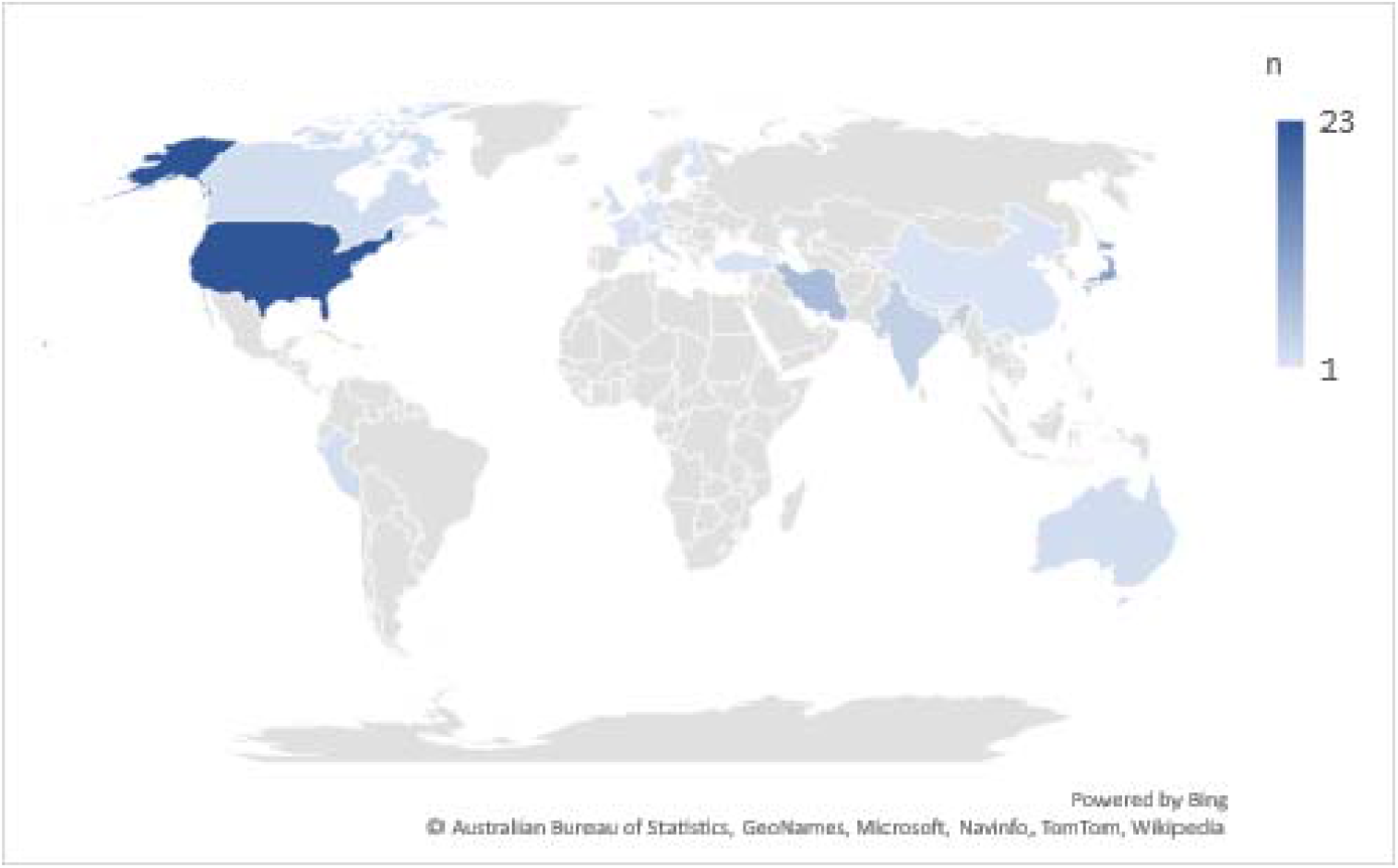
Geographical distribution of articles included * * An article that considered 21 countries in their analysis is not considered in this figure.

### Suicide deaths

Of 40 studies that examined suicide, four were case reports/series ^41-44^, and 11 studies did not conduct any inferential analysis (testing hypotheses statistically) ^30-33,37,38,45-48^. Overall, 15 countries studied, of which seven (including case report study design) were in low- to upper-middle-income countries ^33,37,43,44,49-52^ and the remaining were in high-income countries. One study included data from 21 countries, including 5 low- to upper-middle-income countries and 15 high-income countries ^8^. Findings are grouped based on country income and presented below.

All these studies found an overall no change (n= 4) or a declining trend (n=10) except for studies conducted in Japan in the later months of the pandemic. Japan showed a declining trend for the first three months, but then suicide increased. In Italy, in the two months of February (beginning of pandemic) and April 2020 (the highest COVID-19 daily death), the suicide rate was higher than the rate for the same period in the previous year. Studies (n=15) conducted in Japan ^23,24,36,53-57^, Australia ^58,59^, Austria ^27,60^, US ^61^, Canada ^62^, Germany ^63^, and the study that included 21 countries^8^ used time-series analysis or considered adjusting for the time confounder (i.e. suicide trend) in their analysis.

Low- to upper-middle-income countries included in this review were India ^25,42,48^, Peru ^51,52^, Nepal ^37,50^, China ^49^, Sri Lanka ^33^, Turkey (case-report) ^41^ and Iran (case reports) ^43,44^. Inconsistent findings in two studies from different areas of India were reported. One study from New Delhi showed an initial decline with an increase in the post-lockdown period reaching to pre-COVID-19 rate ^25^. The other study ^48^ reported similar findings to Nepal ^37,50^, with an overall increase in suicide deaths. The study findings from Peru ^51,52^, have a similar pattern as the New Delhi study; China reported an 18% decline ^49^. A study in Sri Lanka assessed self-poisoning (intentional) death rate during COVID-19 compared to pre-pandemic and found a drop in numbers ^33^.

In addition to studies that examined suicide, one study descriptively assessed unnatural deaths over the 6 months of the pandemic and reported that poisoning and hanging cases comprised over 45% percent of the cases at North Bengal in India. Persons involved in private jobs (44%) were more severely affected compared to the government jobs ^64^.

### Overdose deaths

Of 30 studies that assessed overdose and poisoning deaths, five examined unintentional overdose, 10 all intent overdose and some assessed particular substance overdoses e.g. alcohol (n=10). One study was examined accidental poisoning only ^49^. Of these studies, 5 were case report/series or descriptive (i.e. no comparison), and 16 did not conduct any inferential analysis. Only three studies used time-series analysis or considered adjusting for the time trend confounder ^28,65,66^. Studies were conducted in six countries (US, UK, Canada, France, China, and Iran).

There were seven studies that assessed poisoning from ingestion of alcohol-based disinfectant or illicit alcohol (e.g. methanol poisoning). These studies were conducted in US (n=1, case series) ^39^, France (n=1, case report) ^67^ and Iran ^35,68-71^ (n=5). However, the intention of ingestion was not evident in these studies. Particularly in the Islamic country of Iran, where alcoholic drinks are prohibited, it is unclear if the consumption of illicit alcohol is for recreational purposes or due to the spread of misinformation in that country about disinfecting the digestive tract to prevent COVID-19 infection ^68^. Thus, these studies were treated separately. Of these studies, five that compared deaths from methanol poisoning with pre-pandemic figures or previous methanol poisoning outbreaks all showed a considerable increase ^35,68-70^.

Most of the studies that conducted comparison between pre- and post-pandemic periods (n=20) showed that overdose death has been increased (n=15 ^28,29,34,40,65,66,72-81^, 7 with significant findings ^28,29,65,66,72,73,78^) compared to pre-pandemic figures. Only a few studies found null (n=3) ^82-84^ or a decrease in overdose death (n=2) ^49,85^. The study conducted in China evaluating accidental poisoning revealed a decrease compared to previous year’s figures ^49^.

Five studies investigated the type of substance overdosed ^72,73,79,81,82^ or examined a specific drug e.g. methadone (a prescription opioid) ^78,83^ or alcoholic drinks ^78^. Findings from these studies showed that the use of fentanyl (n=4) ^72,73,81,82^, and stimulants such as cocaine and amphetamines ^73,82^ increased significantly compared to the pre-pandemic period. Further, of three studies measuring alcohol overdose deaths, all demonstrated an increase after the pandemic, with one showing the rise being considerably higher than other drug deaths (5.5 folds increase versus 2.5 folds) ^79^. Findings on deaths from heroin overdose were inconsistent, with 1 indicating an increase ^72^ and 2 others showing a decrease ^81,82^. One study found a decline in Benzodiazepines and Fentanyl analogues overdose death ^82^. Studies that examined overdose deaths related to prescription opioids ^81,82^ or overdose deaths among patients receiving treatment for substance misuse ^83,84^ did not observe any change in prescription opioids or overdose deaths among treatment recipients during the pandemic.

### Liver disease

Two studies assessed alcohol-related liver disease or cirrhosis deaths, both conducted in the US ^86,87^. The study that assessed deaths from alcohol-related liver disease (n=1) from a single liver-transplant centre showed a higher number of deaths during the COVID-19’s declining phase compared to the previous year, but this difference was not statistically significant ^86^. UK’s national statistics data found that there is overall an increasing trend of 1.6%. This trend increased even more rapidly and has been statistically higher during COVID-19 up to 4.6% after adjusting for age. This increase was evident in patients in the age group 25–74 years ^87^.

### Inequities

Some studies examined deaths of despair by population characteristics. Eighteen studies assessed the difference in sex and 14 in age groups for suicide death in Japan ^24,47,53,55-57,88^, Australia ^58,59^, US ^61,89^ Peru ^51,52^, Korea ^30^, Norway ^38^, China ^49^, India ^48^ and Italy ^32^. Studies conducted in Japan and Korea showed that the suicide rate significantly increased or showed higher rates among women and younger age groups than men and older age groups. Financial loss in these two groups was higher than other groups. In China, in addition to the younger age group, the elderly group’s suicide rate also increased while the overall trend of suicide was declining compared to the previous year ^49^.

Two studies in the US, assessed changes in the suicide rate among different ethnic groups, reporting an increase among Black and a decrease among White people. Occupations were measured in three studies in Japan, indicating that unemployed homemakers ^56,57^ and students had a higher risk of suicide ^31,56,57^. However, no change was observed among those with recent unemployment in Australia ^59^. A case series study also found a higher number of suicide deaths among daily wagers and self-employed compared to those who worked for the government during the pandemic ^64^. The suicide rate in students showed no change during the school closure in Japan ^54,56^. Studies investigating suicide motives (Japan and Australia; n=2) and suicide methods (Japan and US, n=2) found no differences.

Seven studies assessed changes in overdose-related death based on sex ^65,72,73,75,78,82,87^. Of those, three reported a higher increase in overdose death in men than women ^73,75,78^. Regarding the age group, three studies found no association ^65,72,74^, two observed a rise in younger people (less than 35 years ^73^, 25 years ^34^), and one reported an increase in adults older than 65 years ^34^. 2.6% increase in the average age of overdose deaths is also reported in one study ^82^. One study also found a higher rise in overdose deaths among homeless individuals ^72^. Among studies examining changes in overdose death based on ethnicity (all US; n=5), four reported that the increase was either higher in Black ethnicity or had opposite direction favouring White ethnicity (i.e. Black increased and white decreased).

## Discussion

This rapid review appraised evidence from 70 published studies conducted in 17 countries concerning potential impacts of COVID-19 associated socioeconomic disruption and social isolation on deaths of despair. Findings varied depending on the outcomes. The rate of suicides was not observed to increase during the pandemic ^32,38,45,59,61,90^, though some studies indicated a potential drop in comparison to pre-pandemic years ^26-28,33,46,49,51,52,58,62,89,91^. There were only a few countries, such as Japan, reported contradictory results for suicide. Studies examining overdose death, however, mostly showed a higher rate of overdose death during the COVID-19 period compared to pre-pandemic years. Findings from the studies sub-analysis indicate that women, ethnic minorities and younger age groups, may have suffered disproportionately more than other groups. Note that studies mainly conducted a preliminary data analysis, with several limitations, and the mid-to-longer-term impact of COVID-19 on deaths of despair is to fully emerge.

There were several limitations to the studies that may alter the results. Studies are ecological or cross-sectional in design, mainly had low qualities, and a considerable number of them did not conduct any inferential analysis or only conducted basic comparison without taking account of underlying confounders such as time trends and population on growth. For example, only some of studies used more advanced statistical methods such as interrupted time-series ^8,24,27,28,36,51-54,56-59,61-63,66^ to compare pandemic with pre-pandemic periods. Further 14 studies were either case report or did not use any control group (pre-pandemic period). This could pause a remarkable bias to findings. Data on most recent death cases, particularly in suicide and overdose cases, could be the least reliable and subject to undercounts. As unnatural death case examinations may take an extended amount of time. Further, during the pandemic, the data-collection processes may be disrupted further. Some of the studies were also published as editorials, such as a letter to editor or commentary where they may not have been gone through a peer-review process.

Findings regarding suicide death rates during the pandemic are consistent with a study that analysed data from 21 countries showing either no changes or reductions in suicide ^8^. The lack of increase in suicides since the pandemic began could be attributed to various factors. Despite the early evidence highlighting that health measures such as lockdowns, school closures, and business shutdowns may heighten depression, anxiety, and suicidal thinking ^1^, country policies may have attenuated these adverse effects. Most of the studies have been conducted in high-income countries where welfare safety nets and, in particular, vaccination access, were often greater in comparison with low-income or lower-middle-income countries, which account for 75% ^92^ of the world’s suicides and might have been hit particularly hard by the pandemic. Note that some of these supports, such as financial aids, may now be reduced or halted ^8^. For example, the Australian government had stopped disaster payments to those who have been impacted when 80% of the population became fully vaccinated ^60^ and the observed initial support from the government has faded away over time in Austria ^93^. Thus, it is possible that the pandemic’s potential suicide-related effects are yet to occur even in countries with no current change. This is reflected in some subgroup analyses of the included studies indicating groups who had the highest financial loss or were disadvantaged such as the younger population and women, showed a higher rate of suicide compared to the pre-pandemic period.

Drug overdose and drug-related liver disease deaths, on the other hand, seem to be increased or accelerated remarkably since the pandemic began, particularly in groups subject to iniquity such as Black ethnicity. Our findings regarding overdose deaths are consistent with a previous systematic review conducted on studies and public health surveillance data published prior to Sep 2020 ^5^. In addition, the current review also shows that this higher rise is mainly attributed to synthetic opioids, stimulants, and alcohol overdose deaths which had been observed prior to the pandemic ^82,94^ and have now been accelerated. While these findings are preliminary and limited to a few counties (mostly US), they are concerning and call for urgent actions of policymakers to prevent drug-related deaths rooted in race equity approaches. Strategies such as allowing longer prescription duration, mail, and remote supplying of medications to treat substance use disorders, providing safer drug alternatives such as tablet-based or low release morphine have been suggested as new strategies to reduce harm and drug overdose ^95^

Future studies may want to examine factors such as environmental characteristics that may alleviate or aggravate pandemic-related stress and impacts on deaths of despair. Findings from a survey of 6,080 participants indicated that those who utilized green or blue nature during the pandemic showed a lower score of depression and anxiety and believed that nature helped them cope with COVID-19 stressors ^96,97^. In the current review, none of the included studies has investigated variation in deaths of despair regarding nature exposure during the pandemic. Enabling people to access natural settings on a more frequent basis could be a potential approach to alleviate health inequity through disruption of maladaptive rumination and social anxieties that sustain depression, loneliness and concomitant feelings of despair ^98,99^.

### Conclusion

This rapid review highlights the need for more high-quality studies in general, and in low-middle income countries in particular, to identify the impact of COVID-19 on deaths of despair. Future studies may want to consider the contribution of personal, social, economic and environmental factors that protected some groups while leaving others more vulnerable. Further, despite studies being at a preliminary stage, the change in overdose deaths is concerning and needs strategies to prevent drug overdoses.

## Data Availability

All data produced in the present work are contained in the manuscript.

## Acknowledgment

Ethics approval was not applicable for this systematic review. Funding:. This work was supported by a National Health and Medical Research Council Boosting Dementia Research Leader Fellowship (#1140317) and by a National Health and Medical Research Council Career Development Fellowship (#1148792). All aspects related to the conduct of this study, including the views stated and the decision to publish the findings, are that of the authors only.

## Data availability statement

There is now data collected for this systematic review.

## Data deposition

NA

