## Supplementary materials for "Have deaths of despair risen during the COVID-19 pandemic? A rapid systematic review"

**Table S1. Search strategy**

**EMBASE via Ovid (1947 to 2020) – 29 Aug 2020**

| Despair | 1. | *hopelessness/ |
| --- | --- | --- |
|  | 2. | exp hopelessness/ |
|  | 3. | Despair*.ti,ab,kw. |
|  | 4. | Hopeless*.ti,ab,kw. |
|  | 5. | defeat*.ti,ab,kw. |
|  | 6. | exp social defeat/ |
|  | 7. | *guilt/ |
|  | 8. | guilt*.ti,ab,kw. |
|  | 9. | worthless*.ti,ab,kw. |
|  | 10. | *pessimism/ |
|  | 11. | pessimis*.ti,ab,kw. |
|  | 12. | ((loss or lack or limited) adj3 hope).ti,ab,kw. |
|  | 13. | *anhedonia/ |
|  | 14. | anhedonia*.ti,ab,kw. |
|  | 15. | *apathy/ |
|  | 16. | apath*.ti,ab,kw. |
|  | 17. | self-harm.ti,ab,kw. |
|  | 18. | (drug* or alcohol).ti,ab,kw. |
|  | 19. | *depression/ |
|  | 20. | *anxiety/ |
|  | 21. | depress*.ti,ab,kw. |
|  | 22. | anxiet*.ti,ab,kw. |
|  | 23. | *sadness/ |
|  | 24. | sadness*.ti,ab,kw. |
|  | 25. | *irritability/ |
|  | 26. | *hostility/ |
|  | 27. | (hostilit* or irritabilit*).ti,ab,kw. |
|  | 28. | *loneliness/ |
|  | 29. | (loneliness or lonely).ti,ab,kw. |
|  | 30. | (Loss adj2 self).ti,ab,kw. |
|  | 31. | ((inability or lack or loss) adj3 pleasure).ti,ab,kw. |
|  | 32. | (selfharm* or self-harm*).ti,ab,kw. |
|  | 33. | (opioid* or overdose* or heroin or opium or Fentan?l or methadone or methamphetamine or cocaine or substance abuse or substance misuse or substance addiction).ti,ab,kw. |
|  | 34. | (((economic or financial) adj1 (hardship* or stress* or problem* or cris?s or insecurit* or derpression*)) or recession* or austerity or fiscal cris?s or job loss* or banking cris?s or personnel downsi?ing or macroeconomic condition* or unemployment).ti,ab,kw. |
|  | 35. | ((drug* or alcohol) adj1 (abuse or addiction or misuse or death* or disorder* or dependen*)).ti,ab,kw. |
|  | 36. | ("drug use" or "alcohol use" or alcoholic* or alcoholism).ti,ab,kw. |
| Drug-related liver disease | 37. | (alcoholic liver or toxic liver or chronic hepatitis or (fibrosis adj3 liver) or cirrhosis or poisoning*).ti,ab,kw. |
|  | 38. | *chronic hepatitis/ or alcohol liver cirrhosis/ or "substance use".ti,ab,kw. |
|  | 39. | 1 or 2 or 3 or 4 or 5 or 6 or 7 or 8 or 9 or 10 or 11 or 12 or 13 or 14 or 15 or 16 or 17 or 19 or 20 or 21 or 22 or 23 or 24 or 25 or 26 or 27 or 28 or 29 or 30 or 31 or 32 or 33 or 34 or 35 or 36 or 37 or 38 |
| Death | 40. | *death/ or *fatality/ |
|  | 41. | *mortality/ |
|  | 42. | (fatal* or mortalit* or death*).ti,ab,kw. |
| Suicide | 43. | *suicide/ or suicid*.ti,ab,kw. |
|  | 44. | (selfmutilat* or self-mutilat* or self?mutilation or automutilation or self?immolation or self?immolat* or self?inflict*).ti,ab,kw. |
|  | 45. | ((kill* or hang*) adj1 self).ti,ab,kw. |
|  | 50. | 40 or 41 or 42 |
|  | 51. | 39 and 50 |
|  | 52. | 51 or 42 or 43 or 44 or 45 |
|  | 53. | limit 52 to (human and english language and covid-19) |
|  | 54. | limit 53 to conference abstracts |
|  | 55. | limit 53 to "systematic review" |
|  | 56. | 53 not (54 or 55) |

**Table S2. Study summary and characteristics for suicide outcome**

| **Author year** | **Country, region** | **Study Design** | **Population sample size  %female Age** | **Pandemic start date** | **Pandemic time-period** | **Comparison time-period period/comparison** | **Background information of study pandemic period** | **Comparison** | **Direction (↑↓∅)** |
| --- | --- | --- | --- | --- | --- | --- | --- | --- | --- |
| Anzai 2021 ^(41)^ | Japan, all | ES | General population All F: NA Any age | Mar 2020 | Mar - Jun 2020 | Jan 2013 - Apr 2020 | Lockdown | Before COVID-19 same time | M: ↓*  F: ↓ & then ↑ |
| Barbic 2021 ^(42)^ | Canada, BC | ES | General population All F: NA Any age | Mar 2020 | Mar - Aug 2020 | Jan 2010 - Mar 2020 | First wave of pandemic, economical support provided by government | Period preceding COVID-19 | ↓ |
| Behera 2021 ^(25)^ | India, two districts of New Delhi | ES | General population All F: NA Any age | Mar 2020 | 25 Mar - 31May 2020 | 1 Jun - 31 Oct 2020 | Lockdown | Pandemic post-lockdown period | ↓* (Lockdown lower than post-lockdown) |
|  |  |  |  |  | 25 Mar - 31 May 2020 | 25 Mar - 31May 2019 | Lockdown | Before COVID-19 same time | ↓* |
|  |  |  |  |  | 1 Jun - 31 Oct 2020 | 1 Jun - 31 Oct 2019 | Post-lockdown | Before COVID-19 same time | ∅ |
| Bray 2021 ^(26)^ | US, MD | Es | General population All F: NA Any age | 5 Mar 2020 | 5 Mar - 7 May 2020 | 5 Mar - 7 May, 2017 - 2019 | Lockdown | Before COVID-19 same time | ↓* |
|  |  |  |  |  | 8 May - 7 Jul 2020 | 8 May - 7 Jul, 2017 - 2019 | Post-lockdown | Before COVID-19 same time | ↓* |
| Calati 2021 ^(43)^ | Italy, Milan and Monza and Brianza | ES | Autopsy cases  All F: NA Any age | Mar 2020 | T1: Jan - Apr 2020 | CT1: Jan - Apr 2019 | NR | Before COVID-19 same time | ↓† |
|  |  |  |  |  | Feb-21 | Feb-19 |  |  | ↑ |
|  |  |  |  |  | Mar-21 | Mar-19 |  |  | ↓† |
|  |  |  |  |  | Apr-21 | Apr-19 |  |  | ↑† |
|  |  |  |  |  | 2020 | 2019 |  | Before COVID-19 same time | ↓† |
|  |  |  |  |  | 2020 | 2016 - 2019 |  | Before COVID-19 year by year comparison | ↓† |
| Calderon-Anyosa 2021a ^(44)^ | Peru, all | ES | Adult population All F: NA 18 + y | Mar 2020 | 16 Mar - 30 Jun 2020 | 16 Mar - 30 Jun, 2018 - 2019 | Lockdown | Before COVID-19 same time | ↓(NR) |
|  |  |  |  |  | 1 Jul - 31 Dec 2020 | 1 Jul - 31 Dec, 2018 - 2019 | Post-lockdown | Before COVID-19 same time | ↓(NR) |
| Calderon-Anyosa 2021b ^(45)^ | Peru, all | ES | General population All F: NA Any age | Mar 2020 | 16 Mar - Sep 2020 | 2017 - 1 Mar 2020 | Mar - Jun lockdown Jun -forward easing of restrictions | Pre-lockdown annual slope,  lockdown monthly difference,  pre-lockdown slope interaction | M: ↓(NR); An increase in time trend slope (1.2 more deaths/million after lockdown compared to the pre-pandemic period  F: ↓(NR); no change in time trend slope |
| Deisenhammer 2021 ^(27)^ | Austria, state of Tyrol | ES | General population  All  F: NA  Any age | NR | 1 Apr - 30 Sep 2020 | 2 Apr - 30 Sep, 2006 - 2019 | NR (includes lockdown) | Before COVID-19 same time | ↓* |
|  |  |  |  |  | 1 Apr - Jun 2020 | 1 Apr - Jun, 2006-2019 | NR | Before COVID-19 same time | ↓ |
|  |  |  |  |  | Jun - 30 Sep 2020 | Jun - 30 Sep, 2006 - 2019 | NR | Before COVID-19 same time | ↓* |
| Dwyer 2021 ^(46)^ | Australia, VIC | ES | General population All F: NA Any age | 27 Feb 2020 | Mar 2020 - Jan 2021 | 2015 - 2019 | Lockdown and Post-lockdown periods | Before COVID-19 same time | ↓* |
| Faust 2021b ^(47)^ | US, MA | ES | General population  All F: NA 10 + y | Mar 2020 | Mar - May 2020 | Jan - May, 2015 - 2019 | Lockdown | Before COVID-19 same time | ∅ |
| Faust 2021a ^(47)^ | US, all | ES | General population All F: NA Any age | Mar 2020 | Mar - Aug 2020 | Mar - Aug, 2015 - 2019 | NR (includes lockdown) | Before COVID-19 same time | ↓* |
| Habu 2021 ^(48)^ | Japan, two cities of Okayama and Kibichuo | ES | General population All F: NA Any age | Mar 2020 | Mar - Aug 2020 | Mar - Aug, 2018 - 2019 | Lockdown and post-lockdown | Before COVID-19 same time | ∅ |
| Isumi 2020 ^(49)^ | Japan, All | ES | Children All F: NA < 20 y | Mar 2020 | Mar - May 2020 | Mar - May, 2018 - 2019 | School closure | Before COVID-19 same time | ∅ |
| Karakasi 2021 ^(29)^ | Greece, regional unit of Evros, Thrace | ES | General population All F: NA Any age | Feb 2020 | 1 Mar - 15 May 2020 | 1 Mar - 15 May, 2010 - 2019 | Lockdown | Before COVID-19 same time | ∅ |
| Nomura 2021a and Nomura 2021b (update) ^(23, 24)^ | Japan, all | ES | General population All F: NA Any age | Mar 2020 | Mar – Dec 2020 | Mar - Sep, 2016 - 2019 | Lockdown (April-May) | Before COVID-19 same time (expected mortality) | M: Jan & Oct: 2-22% ↑*; Apr 4-14% ↓*  F: Feb & Apr: 1-18% ↓*; Jul-Dec (Post-lockdown): 21-85%↑* |
| Kim 2021b ^(30)^ | Korea, all | ES | General population All F: NA Any age | Jan 2020 | Jan - Aug 2020 | Jan - Aug 2019 | Mar highest number of COVID-19 | Before COVID-19 same time | Total: ↓†  M: ↓  F: ↑ |
| Leske 2021 ^(50)^ | Australia, QLD | ES | General population All F: NA Any age | Mar - 2021 (restrictions), Feb covid arrival | Feb - Aug 2020 | 2015 - Jan 2020 | Lockdown and Post-lockdown | Before COVID-19 same time | ∅ |
| Marutani 2021 ^(31)^ | Japan, all | ES | University graduate students All F: 30 Any age | Apr 2020 | Apr 2020 - Mar 2021 | 2002 - 2019 | Courses all online | Before COVID-19 same time | ↑† |
| Messina 2021 ^(32)^ | Italy, all | ES | Italian Police All F: NA 25 + y | Feb 2020 | 2020 | 2015-2019 | Lockdown: Feb - Aug 2020 | Before COVID-19 annual average | ∅ |
| Mitchell 2021 ^(51)^ | US, CT | ES | General population All F: NA Any age | Mar 2020 | Mar - May 2020 | Mar - May 2014-2019 | Lockdown | Before COVID-19 same time | Lockdown vs 5 y average: 20% ↓†  Lockdown vs previous y: 13% ↓† |
| Ontiveros 2021§ ^(52)^ | US, CA | CS | Suicide cases reported to a poison centre  All F: NA Any age | Mar 2020 | Mar - May 2020 | Mar - May, 2018-2019 | Lockdown | Before COVID-19 same time | ↓ |
| Osaki 2021 ^(53)^ | Japan, all | ES | General population All F: NA Any age | Apr 2020 | Mar - Dec 2020 | Average of 3 y prior to covid | Lockdown and post-lockdown | Before COVID-19 (3 y average) | 1st wave (Apr-May): overall: 18%↓*  2nd wave: F: ↑ *, M: ∅  3rd wave: overall: ↑*, F: 70% ↑* |
| Pirkis 2021 ^(8)^ | 21 countries (16 high-income and 5 upper-middle-income) | ES | General population All F: NA Any age | NR | 1 Apr - 31 Jul 2020 | Jan 2019 - Mar 2020 | Lockdown and post-lockdown | Period preceding COVID-19 | ∅ or ↓ |
| Pokhrel 2021 ^(37)^ | Nepal, all | ES | General population All F: NA Any age | Mar 2020 | Mar - Jun 2020 | NR (pre-lockdown date not specified) | Lockdown | Before covid (period NR) | 25% ↑† |
| Qin 2021 ^(38)^ | Norway, all | ES | General population All F: NA Any age | Mar 2020 | Mar - May 2020 | Mar - May, 2014-2018 | Lockdown | Before COVID-19 same time | ∅ |
| Radeloff 2021 ^(54)^ | Germany, Leipzig major city | ES | General population All F: NA Any age | Mar 2020 | T1: 17- 22 Mar 2020 & 6 Jun - 30 Sep 2020 | CT1: Jan - 17 Mar 2020 | T1: travel restriction (moderate) | Period preceding COVID-19 | ∅ |
|  |  |  |  |  | T2: 22 Mar - 5 Jun 2020 | CT1: Jan - 17 Mar 2020 | T2: travel, outing and socialising restriction (severe) | Period preceding COVID-19 | T2 vs CT1: ↓*  T2 vs T1: ∅ |
|  |  |  |  |  | T overall: Mar - Sep 2020 | CT overall: Mar - Sep 2010- 2019 | Lockdown and post-lockdown | Before COVID-19 same time | ∅ |
| Sakamoto 2021 ^(55)^ | Japan, all | ES | General population  All  F: NA  Any age | Central and local Gov: Late Mar 20 20 , National: Apr 2020 | Apr - Nov 2020 | Apr – Nov, 2016 - 2019 | Lockdown (until May) and post-lockdown | Before COVID-19 same time | M: Oct - Nov: ↑*  F: Jul - Nov: ↑* |
| Sengupta 2020 ^(56)^ | India, Cooch Behar (4th most populous state) | CS | Autopsy cases All (n=335) F: NA Any age | 25 Mar 2020 | 25 Mar - 25 Apr 2020 | Apr 2019 Jan 2020 Feb 2020 Mar 2020 | Lockdown | Period preceding COVID-19 and before COVID-19 same time | ↑ † regardless of comparison period |
| Seposo 2021 ^(36)^ | Japan, all | ES | General population All F: NA Any age | Apr 2020 | Apr - Dec 2020 | Jan 2010 - Mar 2020 | Lockdown: Apr - May 2020 and post-lockdown | Before COVID-19 (10 y average) | Relative risk change: ↑*  Crude comparison: ↓* |
| Shrestha 2021 ^(57)^ | Nepal, Kathmandu (Dhulikhel rural tertiary) | CS | Patients with self-harm at an emergency department All F: NA Any age | 24 Mar 2020 | 24 Mar - 23 Jun 2020 | 25 Mar-23 Jun 2019 | Lockdown | Period preceding COVID-19 | ↑† |
|  |  |  |  |  |  | 24 Dec 2019 - 23 Mar 2020 |  | Period preceding COVID-19 | ↑† |
| Tanaka 2021 ^(58)^ | Japan, all | ES | General population All (126M) F: NA Any age | NR | Outbreak 1: Feb - Jun 2020 | Nov 2016 - Feb 2020 | 1^st^ Outbreak | Before COVID-19 | 14% ↓* |
|  |  |  |  |  | outbreak 2: Jul - Oct 2020 |  | 2^nd^ outbreak |  | 16% ↑* |
| Ueda 2021 ^(59)^ | Japan, all | ES | General population  All  F: NA  Any age | NR | Apr - Oct 2020 | 2017- Feb 2019 | Two outbreaks | Before COVID-19 trends (3 y) | Feb - Jun: ↓*  Jul - Oct: ↑* |
| Zheng 2021^(60)^ | China, Guangdong | ES | General population All F: NA Any age | Jan 2020 | 1 Jan - 30 Jun 2020 | 1 Jan - 30 Jun 2019 | Lockdown and post-lockdown | Before COVID-19 same time | Overall: (-)18.46% ↓*  0-14 y: 139.2% ↑*  70-79 y: 16.8% ↑* |
| Carlin 2021 ^(61)^ | Austria, Vienna | CS | A trauma centre admission cases All F: NA Any age | 16 Mar 2020 | 16 Mar - 15 May 2020 | 16 Mar - 15 May, 2015- 2019 (except for 2017 as the hospital was closed) | Lockdown | Before COVID-19 same time | ∅ |
| Knipe 2021§ ^(33)^ | Sri Lanka, | CS | Patients presented with self-poisoning All F: NA Any age | 20 Mar 20 | 20 Mar - 31 Aug 2020 | 1 Jan 2019 - 19 Mar 2020 | Lockdown and post-lockdown (lockdown lifted in 28 Jun 2020) | Period preceding COVID-19 | ↓† |
| Sakelliadis 2020 ^(62)^ | Greece, Athens | CS | Autopsy cases All F: NA Any age | 17 Mar 20 | 17 Mar - 15 Apr 2020 | 17 Mar - 15 Apr 2019 | Lockdown | Before COVID-19 same time | ∅ |
| Kumar 2021^‡ (63)^ | India, West Bengal | CS | Autopsy cases  All  F: NA  Any age | Mar 2020 | Mar - Sep 2020 | None | Lockdown | NA | 27.8% and 17.9% of autopsy cases were related to hanging and poisoning, respectively. |
| Choudhury 2020 ^(64)^ | India, Lucknow | CSS | Suicide cases All (n=59) F: 44.4% Any age | Mar-20 | 24 Mar-31May 2020 | NA | Lockdown | NA | Economic factors accounted for 49% of cases, domestic conflicts 23.7%, psychological and emotional factors 27.1%.  64% of suicides were in the group of 18 -35 y.  The majority of suicides were done by hanging (93.2%).  The daily wagers and the self-employed citizens were most affected (36%). |
| Forouzanfar 2020 ^(65)^ | Iran, Tehran | CR | Family suicide A middle-aged mother and a 32 y son (n=2) | NR | NR | NA | NR | NA | The reason for suicide was the death of the father due to COVID-19  Method: poisoning (aluminium phosphide)  Background: wealthy and highly educated, high Socioeconomic status |
| Pirnia 2020 ^(66)^ | Iran, Tehran | CR | Family suicide A teenager son and 52 y mother (n=2) | NR | 18-20 Mar 2020 | NA | Lockdown | NA | Father died due to COVID-19 few weeks back. Lack of mourning rites specified as a reason for his depression symptoms and suicide.  Mother committed suicide by taking 2 aluminium phosphide pills two days after his son death. |
| Uğurlu 2020 ^(67)^ | Turkey | CR | Suicide case 34 y male | NR | 02-Apr-20 | NA | NR | NA | Reason: Mixed anxiety and depression induced by the COVID-19 pandemic stressor  Method: gun shot at his home.  Background: no know history of psychiatric or physiological disorder, no regular job |

§ Suicide due to poisoning only, ^‡^ hanging and poisoning

† No inferential comparison

↑ denotes increase; ↓denotes decrease; ∅ no considerable change; * p<0.05

CS cross-sectional study; CT control time; CSS case series study; CR case-report study; ES ecological study; F female; M male, NA not applicable; T time

**Table S3. Study summary and characteristics for overdose death and drug-related liver disease death**

| **Author Year** | **Country, region** | **Study Design** | **Population sample size  %female Age** | **Pandemic start date** | **Pandemic time-period** | **Comparison time-period period** | **Background information** | **Death outcome** | **Comparison** | **Direction (↑↓**∅**)** |
| --- | --- | --- | --- | --- | --- | --- | --- | --- | --- | --- |
| Appa 2021 ^(68)^ | US, San Francisco | ES | General population All F: NA Any age | Mar 2020 | 17 Mar - 30 Nov 2020 | 1 Jul 2019 - 16 Mar 2020 | NR | Drug overdose (unintentional) | Period preceding COVID-19 | ↑* |
|  |  |  |  |  | 2020 | 2017 - 2019 |  |  | Before COVID-19 (monthly average) | ↑† |
| Brothers 2021 ^(69)^ | US, CT | CSS | Methadone recipients All F: NA Any age | 16 Mar 2020 | Apr - Aug 2020 | Apr – Aug, 2015 - 2019 | Methadone administration has relaxed | Methadone overdose | Before COVID-19 same time | ∅ (no change compared to other opioids’ trends) |
| DiGennaro 2021 ^(70)^ | US, MA | ES | General population All F: NA Any age | Mar 2020 | 24 Mar - 8 Nov 2020 | 24 Mar - 8 Nov 2019  24 Mar - 8 Nov 2018 | Lockdown | Drug overdose (all intent) | Before COVID-19 same time | ∅  ∅ |
| Faust 2021a ^(28)^ | US, all | ES | General population All F: NA Any age | Mar 2020 | Mar - Aug 2020 | Mar - Aug, 2015 - 2019 | NR (includes lockdown) | Drug overdose (all intent) | Before COVID-19 same time | ↑* |
| Friedman 2021 ^(71)^ | US, all | ES | General population All F: NA Any age | Mar 2020 | Jan - Jul 2020 | Jan - Jul, 2015 - 2019 | NR (includes lockdown) | Drug overdose (all intent) | Before COVID-19 same time | ↑† |
| Aghababaeian 2020  ^(72)^ | Iran, All | ES | General population All F: NA Any age | Feb 2020 | 7 Mar - 8 Apr 2020 | Mar - Apr 2019 | An increase in the availability of illegal alcohol-based products | Alcohol (Methanol) | Before COVID-19 same time | ↑† |
|  |  |  |  |  |  | Apr 2016 - Sep 2018 |  |  | Before COVID-19 overall time (annual average | ↑† |
| Karakasi 2021 ^(29)^ | Greece, regional unit of Evros, Thrace | ES | General population All F: NA Any age | Feb 2020 | 1 Mar - 15 May 2020 | 1 Mar - 15 May 2010 - 2019 | Lockdown | Unintentional poisoning | Before COVID-19 same time | ↑* |
| Khatri 2021 ^(73)^ | US, Philadelphia | ES | General population All F: NA Any age | Mar 2020 | Mar - May 2020 | Mar - May 2019 | Lockdown | Opioid overdose (unintentional) | Before COVID-19 same time | Black: ↑*  Non-Hispanic White: ↓  Hispanic: ∅ |
|  |  |  |  |  |  | Dec 2019 - Feb 2020 |  |  | Period preceding COVID-19 | Black: ↑*  Non-Hispanic White: ∅  Hispanic: ∅ |
| Kim 2021a ^(74)^ | US, all | ES | General population All F: NA Any age | Mar 2020 | Q1 - Q3 2020 | Q1 - Q4, 2017 - 2019 | NR | Chronic Liver Disease and Cirrhosis (combined) | Before COVID-19 same time | 4.6% ↑* (age-adjusted) |
| Kitchen 2021 ^(75)^ | Canada, Ontario | ES | General population All F: NA 15 + y | 15 Mar 2020 | 15 Mar - Sep 2020 | 15 Mar - Sep 2019 | Lockdown | Opioid overdoses (all intent) | Period preceding COVID-19 | 135% ↑* |
|  |  |  |  |  |  | Sep 2019 - 15 Mar 2020 |  |  | Before COVID-19 same time | 135% ↑* |
| Hassanian-Moghaddam 2020 ^(76)^ | Iran, all | ES | General population All F: NA Any age | Feb 2020 | Feb - May 2020 | None (outbreak in Libya) | An increase in the availability of illegal alcohol | Alcohol (Methanol) | None (Second-largest methanol outbreak in history) | ↑† |
| Mariottini 2021 ^(77)^ | Finland, all | CS | Autopsy cases  All  F: NA Any age | Mar 2020 | Jan - Aug 2020 | 2015 - 2019 | lockdown and post-lockdown (Jun), reduced  access to harm-reduction services | 3 most common drugs | Before COVID-19 same time | ↑† |
| Mason 2021b ^(40)^ | US, Cook  County | ES | General population All F: NA Any age | Mar 2020 | 21 Mar - 30 May 2020 | CT1: 15 Dec 2019 – 20 Mar 2020  CT2: 1 Jan 2018 - 14 Dec 2019 | Lockdown, interruptions and changes in the illicit drug supply | Opioid overdose (all intent) | Period preceding COVID-19 (average) | ↑†  ↑† |
|  |  |  |  |  | 31 May - 6 Oct 2020 | CT1: 15 Dec 2019 – 20 Mar 2020  CT2: 1 Jan 2018 - 14 Dec 2019 | Post -lockdown |  | Period preceding COVID-19 (average) | ↑†  ∅ |
| Mason 2021a ^(78)^ | US, Cook County | ES | General population All F: NA Any age | Mar 2020 | 21 Mar - 30 May 2020 | CT1: 15 Dec 2019 – 20 Mar 2020  CT2: 1 Jan 2018 - 14 Dec 2019 | Lockdown, interruptions in the illicit drug supply and in-person service | Opioid overdose (all intent) | Period preceding COVID-19 (average) | Lockdown vs CT1 & CT2: ↑ |
|  |  |  |  |  | 6 Jun - 23 Dec 2020 |  | Post-lockdown, interruptions in the illicit drug supply and in-person service |  |  | Post-lockdown vs CT1 & CT2: ↑  Lockdown vs post-lockdown: ↑ |
| Shokoohi 2020 ^(79)^ | Iran, all | CS | General population All F: NA Any age | Feb 2020 | Feb - Apr 2020 | Feb - Apr 2019 | An increase in the availability of illegal alcohol-based products | Alcohol (Methanol) | Before COVID-19 same time | ↑† |
|  |  |  |  |  |  | Mar 2016 - Aug 2018 (29 months) |  |  | Before COVID-19 (29 months) | ↑† |
| Patel 2021 ^(80)^ | Us, Birmingham | CS | Patients admitted for an overdose One hospital’s patients F: NR Any age | NR | 1 Jan - 31 Oct 2020 | 1 Jan - 31 Oct, 2019 | Lockdown and post-lockdown | Opioid overdose (all intent) | Before COVID-19 same time | Overall ↓†  Black ~20%↑†  White ~30%↓† |
| Rutledge 2021 ^(81)^ | US, NY | CS | Alcohol-associated liver disease (ALD) patients  All  ALD patients referred to a liver transplantation centre  F: 37-50%  Any age | Mar 2020 | T2: 23 Apr -23 Aug 2020 | 1 Jan - 21 Mar 2020 | Post-lockdown | ALD | Period preceding COVID-19 | ↑ |
| Shreffler 2021 ^(82)^ | US, KY (Jefferson County, most populated) | CS | General population All F: NA Any age | 6 Mar 2020 | 6 Mar - 25 Jun 2019 | 6 Mar - 25 Jun 2019 | Lockdown | Drug overdose (all intent) | Before COVID-19 same time | ↑† |
|  |  |  |  |  |  | 15 Nov 2019 - 5 Mar 2020 |  |  | Period preceding COVID-19 | ↑† |
| Vieson 2021 ^(83)^ | US, OH | ES | Adult population  All  F: NA  18 + y | NR | Apr - Jun 2020 | T1: Expected value for 2020  T2: last peak Q4 2017 | NR | Opioid overdose (all intent) | Before COVID-19, expected mortality | ↑*  null |
| Rodda 2020 ^(84)^ | US, San Francisco | ES | General population All F: NA 15 + y, Median: 54 y (IQR: 35-56) | 15 Mar 2020 | 16 Mar - 18 Apr, 2020 | 1 Jan - 15 Mar 2020 | Lockdown | Opioid overdose (accidental) | Period preceding COVID-19 and before COVID-19 same time | ↑† |
|  |  |  |  |  |  | 16 Mar - 18 Apr, 2018- 2019 |  |  |  | ↑† |
| Yazdi-Feyzabadi 2021 ^(35)^ | Iran, 13 provinces | ES | General population All F: NA | Feb 2020 | Apr-20 | None | An increase in the availability of illegal alcohol-based products | Alcohol (Methanol) | None | A high number of poisonings |
| Zhang 2021 ^(34)^ | US, OH | ES | General population All F: NA Any age | 15 Mar 2020 | Mar - 10 Oct 2020 | 1 Jan 2018 - 15 Mar 2020 |  | Drug Overdose (all intent) | Period preceding COVID-19 | Up to 76.8% ↑†  then dropped to an earlier rate when unemployment reduced. |
| Zheng 2021 ^(60)^ | China, Guangdong | ES | General population All F: NA Any age | Jan 2020 | 1 Jan - 30 Jun 2020 | 1 Jan - 30 Jun 2019 | Lockdown and post-lockdown | Poisoning (accidental) | Before COVID-19 same time | ↓† |
| Pines 2021 ^(85)^ | US, 18 states | ES | Overdose visits to EDs  All EDs  F: NA  Any age | 13 Mar 2021 | 13 Mar - 31 Jul 2020 | 14 Mar- 31 Jul 2019 | NR | Drug overdose death (ED deaths or deaths on arrival, all intent) | Before COVID-19 same time | Total: 2.51 times  ↑†  Opioid: 2.5 times ↑†  Alcohol:5.25 times ↑†  Other drugs: 2 times ↑† |
| Slavova 2021 ^(86)^ | US, KY | ES | Kentucky State Ambulance cases All F: NA Any age | 5 Mar 2020 | 6 Mar - 26 Apr 2020 | January 14, 2020, to March 5, 2020 | Lockdown | Opioid overdose death at the scene (all intent) | Period preceding COVID-19 | ↑† |
| Glober 2020 ^(87)^ | US, Marion county | ES | General population All F: NA | 25 Mar 2020 | 25 Mar - 7 July 2020 | 26 Mar -7 July 2019 | Lockdown and post-lockdown (early May) | Any drug overdose (including suspected cases) | Before COVID-19 same time | ↑* |
|  |  |  |  |  |  | 1 Jan 2019 - 24 Mar 2020 |  |  | Period preceding COVID-19 | ↑* |
| Congdon 2021 ^(88)^ | UK, London | CS | Patients registered at two substance misuse services  All  F: NA  Any age | Mar 2020 | Mar - Apr 2020 | Mar - Apr 2019 | Reduced face-to-face interactions in the service | Drug overdose (accidental) | Before COVID-19 same time | ∅ |
| UK National Statistical Bulletin 2021 ^(89)^ | UK, England, and Wales | ES | General population All F: NA | NR | Q1 - Q4 2020 | 2001-2019 | lockdown and post-lockdown | Alcohol | Before COVID-19 same time | 2020 vs 2019: 19.6% ↑*  Q2 - Q4 2020 vs 2001 - Q1 2020: 17-28% ↑*  Continuous since Q2: ↑* trend  2020  Q1 2020 vs 5 y average: ∅ |
| Yip 2020 ^(39)^ | US, Arizona, and New Mexico | CSS | Methanol poisoning cases All F: NA Any age | NR | 1 May - 30 Jun 2020 | NA | NR | Alcohol (Methanol) | None | 4 out of 15 poisoning cases were related to methanol |
| Simani 2020 ^(90)^ | Iran, Tehran | CSS | Methanol poisoning cases underwent computed tomography scan n=40 F: 23% (non-survival) Age: 42.4 ± 14.0 y (non-survival) | NR | Mar - Apr 2020 | NA | NR | Alcohol (Methanol) | None | Mortality rate: 55% (n=22)  The lower mortality rate in cases with chronic alcohol consumption than those who drank alcohol for the first time (p<0.05).  No significant difference in the blood level of Amphetamine, history of other illegal drugs, or COVID-19 infection was found between survivals and non-survivals.  Non-survival characteristics:  Alcohol history: 18 (81.8%) Amphetamine level: 6 (27.3%) Illicit drug history: 10 (45.5%) |
| Dumollard 2021 ^(91)^ | France | CR | Death case due to Isopropyl Alcohol poisoning (n=1) F: 0% 33 y | NR | NR | NA | NR | Alcohol (isopropyl and acetone) (cleaning solvent) | NA | Found dead in his home with a bottle of isopropyl alcohol liquid found close to him  Background: history of drug addiction, psychosis, and an attempt of hanging himself. |

† No inferential comparison

↑ denotes increase; ↓denotes decrease; ∅ no considerable change; * p<0.05

ALD Alcohol-associated liver disease; CS cross-sectional study; CT control time; ED emergency department; ES ecological study; F female; IQR interquartile range; M male, NA not applicable; NR not reported; Q quartile; T time

**Table S4. Quality assessment of included publications (n=70)**

a) **Cross-sectional studies**

For the list of questions assessed for each type of studies, see <https://www.nhlbi.nih.gov/health-topics/study-quality-assessment-tools>

For ecological studies, the adapted questions are:

Q14 (modified): Were key potential confounding variables measured at the ecological unit level and adjusted statistically for their impact on the relationship between exposure(s) and outcome(s)?

Q15 (added): Was spatial autocorrelation addressed?

Q16 (added): Was variation of outcome distribution within each unit of analysis accounted for in any way?

| **Author Year** | **Q1** | **Q2** | **Q3** | **Q4** | **Q5** | **Q6** | **Q7** | **Q8** | **Q9** | **Q10** | **Q11** | **Q12** | **Q13** | **Q14** | **Q15** | **Q16** | **Total score** | **Total applicable** | **Ratio** |
| --- | --- | --- | --- | --- | --- | --- | --- | --- | --- | --- | --- | --- | --- | --- | --- | --- | --- | --- | --- |
| Anzai 2021 | Y | Y | Y | N | NA | N | N | N | Y | N | Y | Y | NA | Y | N | N | 7 | 14 | 0.50 |
| Appa 2021 | Y | Y | Y | N | NA | N | N | N | Y | N | Y | Y | NA | N | N | N | 6 | 14 | 0.43 |
| Barbic 2021 | Y | Y | Y | N | NA | N | N | N | Y | N | Y | Y | NA | Y | Y | N | 8 | 14 | 0.57 |
| Behera 2021 | Y | Y | Y | N | NA | N | N | N | Y | N | Y | Y | NA | N | N | N | 6 | 14 | 0.43 |
| Bray 2021 | Y | Y | Y | N | NA | N | N | Y | Y | Y | Y | Y | NA | N | N | N | 8 | 14 | 0.57 |
| Brothers 2021 | Y | Y | Y | N | NR | N | N | N | Y | N | Y | Y | NA | N | NA | NA | 6 | 13 | 0.46 |
| Calati 2021 | Y | Y | Y | N | NA | N | N | N | N | N | Y | Y | NA | N | N | N | 5 | 14 | 0.36 |
| Calderon-Anyosa 2021a | Y | Y | Y | N | NA | N | N | Y | Y | Y | Y | Y | NA | N | NR | NR | 8 | 14 | 0.57 |
| Calderon-Anyosa 2021b | Y | Y | Y | N | NA | N | N | Y | Y | Y | Y | Y | NA | Y | Y | Y | 11 | 14 | 0.79 |
| Deisenhammer 2021 | Y | Y | Y | N | NA | N | N | Y | Y | Y | Y | Y | NA | Y | NR | NR | 9 | 14 | 0.64 |
| DiGennaro 2021 | Y | Y | Y | N | NA | N | N | N | Y | N | Y | Y | NA | N | NR | NR | 6 | 14 | 0.43 |
| Dwyer 2021 | Y | Y | Y | N | NA | N | N | N | Y | N | Y | Y | NA | Y | Y | NA | 8 | 13 | 0.62 |
| Faust 2021b | Y | Y | Y | N | NA | N | N | N | Y | N | Y | Y | NA | Y | Y | NR | 8 | 14 | 0.57 |
| Faust 2021a | Y | Y | Y | N | NA | N | N | N | Y | N | Y | Y | NA | Y | Y | NR | 8 | 14 | 0.57 |
| Friedman 2021 | Y |  | Y | N | NA | N | N | N | N | N | Y | Y | NA | N | N | N | 4 | 14 | 0.29 |
| Habu 2021 | Y | Y | Y | N | NA | N | N | N | Y | N | N | Y | NA | N | NR | NR | 5 | 14 | 0.36 |
| Hamdanieh 2020 | Y | Y | Y | N | NA | N | N | N | Y | N | Y | Y | NA | N | NR | NR | 6 | 14 | 0.43 |
| Isumi 2020 | Y | Y | Y | N | NA | N | N | N | Y | N | Y | Y | NA | N | NR | NR | 6 | 14 | 0.43 |
| Karakasi 2021 | Y | Y | Y | N | NA | N | N | N | Y | N | NR | Y | NA | N | NR | NR | 5 | 14 | 0.36 |
| Nomura 2021 | Y | Y | Y | N | NA | N | N | N | Y | N | Y | Y | NA | Y | NR | NR | 7 | 14 | 0.50 |
| Nomura 2021b | Y | Y | Y | N | NA | N | N | N | Y | N | Y | Y | NA | Y | NR | NR | 7 | 14 | 0.50 |
| Khatri 2021 | Y | Y | Y | N | NA | N | N | N | Y | N | Y | Y | NA | N | N | N | 6 | 14 | 0.43 |
| Kim 2021 | Y | Y | Y | N | NA | N | N | N | Y | N | Y | Y | NA | N | N | N | 6 | 14 | 0.43 |
| Kim 2021 | Y | Y | Y | N | NA | N | N | N | N | N | Y | Y | NA | Y | N | N | 6 | 14 | 0.43 |
| Kitchen 2021 | Y | Y | Y | N | NA | N | N | N | Y | N | Y | Y | NA | N | N | N | 6 | 14 | 0.43 |
| Hassanian-Moghaddam 2020 | Y | Y | Y | N | NA | N | N | N | Y | N | Y | Y | NA | N | N | N | 6 | 14 | 0.43 |
| Kumar 2021 | Y | Y | Y | Y | NA | N | N | N | Y | N | Y | Y | NA | N | N | N | 7 | 14 | 0.50 |
| Leske 2021 | Y | Y | Y | N | NA | N | N | N | Y | N | Y | Y | NA | N | Y | NR | 7 | 14 | 0.50 |
| Mariottini 2021 | Y | Y | Y | Y | NA | N | N | N | N | N | Y | Y | NA | N | N | N | 6 | 14 | 0.43 |
| Marutani 2021 | Y | Y | Y | N | NR | N | N | N | Y | N | N | NR | Y | N | N | N | 5 | 16 | 0.31 |
| Mason 2021b | Y | Y | Y | N | NA | N | N | N | Y | N | Y | Y | NA | N | N | N | 6 | 14 | 0.43 |
| Mason 2021b | Y | Y | Y | N | NA | N | N | Y | Y | Y | Y | Y | NA | N | N | N | 8 | 14 | 0.57 |
| Messina 2021 | Y | Y | Y | N | NA | N | N | N | N | N | NR | Y | NA | N | N | N | 4 | 14 | 0.29 |
| Mitchell 2021 | Y | Y | Y | N | NA | N | N | N | Y | N | Y | Y | NA | N | N | N | 6 | 14 | 0.43 |
| Shokohi 2020 | Y | Y | Y | N | NA | N | N | N | Y | N | Y | Y | NA | N | N | N | 6 | 14 | 0.43 |
| Ontiveros 2021 | Y | Y | Y | N | NA | N | N | N | Y | N | N | Y | NA | N | N | N | 5 | 14 | 0.36 |
| Osaki 2021 | Y | Y | Y | N | NA | N | N | N | Y | N | Y | Y | NA | N | N | N | 6 | 14 | 0.43 |
| Patel 2021 | Y | Y | Y | N | NA | N | N | N | NR | N | Y | Y | NA | N | NR | NR | 5 | 14 | 0.36 |
| Pirkis 2021 | Y | Y | Y | N | NA | N | N | N | Y | N | Y | Y | NA | Y | NR | NR | 7 | 14 | 0.50 |
| Pokhrel 2021 | Y | Y | Y | N | NA | N | N | N | Y | N | Y | Y | NA | N | NR | NR | 6 | 14 | 0.43 |
| Qin 2021 | Y | Y | Y | N | NA | N | N | N | Y | N | Y | Y | NA | N | NR | NR | 6 | 14 | 0.43 |
| Radeloff 2021 | Y | Y | Y | N | NA | N | N | Y | Y | Y | Y | Y | NA | N | NR | NR | 8 | 14 | 0.57 |
| Rutledge 2021 | Y | Y | Y | Y | NA | Y | N | Y | Y | Y | Y | Y | Y | N | NR | NR | 12 | 15 | 0.80 |
| Sakamoto 2021 | Y | Y | Y | N | NA | N | N | N | Y | N | Y | Y | NA | N | NR | NR | 6 | 14 | 0.43 |
| Sengupta 2020 | Y | Y | Y | Y | NR | N | N | N | Y | N | Y | Y | NA | N | N | N | 7 | 15 | 0.47 |
| Seposo 2021 | Y | Y | Y | N | NA | N | N | N | Y | N | Y | Y | NA | Y | NR | NR | 7 | 14 | 0.50 |
| Shreffler 2021 | Y | Y | Y | Y | NR | N | N | N | Y | N | Y | Y | NA | N | NR | NR | 7 | 15 | 0.47 |
| Shrestha 2021 | Y | Y | Y | Y | NR | N | N | N | Y | N | Y | Y | NA | N | N | N | 7 | 15 | 0.47 |
| Tanaka 2021 | Y | Y | Y | N | NA | N | N | Y | Y | Y | Y | Y | NA | Y | NR | NR | 9 | 13 | 0.69 |
| Ueda 2021 | Y | Y | Y | N | NA | N | N | N | Y | Y | Y | Y | NA | Y | NR | NR | 8 | 13 | 0.62 |
| Vieson 2021 | Y | Y | Y | N | NA | N | N | N | Y | Y | Y | Y | NA | Y | NR | NR | 8 | 13 | 0.62 |
| Rodda 2020 | Y | Y | Y | N | NA | N | N | N | Y | N | Y | Y | NA | N | N | N | 6 | 14 | 0.43 |
| Yazdi-Feyzabadi 2021 | Y | Y | Y | N | NA | N | N | N | Y | N | Y | Y | NA | N | N | N | 6 | 14 | 0.43 |
| Zhang 2021 | Y | Y | Y | N | NA | N | N | N | Y | N | Y | Y | NA | N | N | N | 6 | 14 | 0.43 |
| Zheng 2021 | Y | Y | Y | N | NA | N | N | N | Y | N | Y | Y | NA | N | N | N | 6 | 14 | 0.43 |
| Pines 2021 | Y | Y | Y | N | NA | N | N | N | Y | N | Y | Y | NA | N | N | N | 6 | 14 | 0.43 |
| Carlin 2021 | Y | Y | Y | Y | NR | N | N | N | Y | N | Y | Y | NA | N | NA | NA | 7 | 13 | 0.54 |
| Slavova 2021 | Y | Y | Y | N | NA | N | N | N | Y | N | Y | Y | NA | N | N | N | 6 | 14 | 0.43 |
| Glober 2020 | Y | Y | Y | N | NA | N | N | N | Y | N | Y | Y | NA | Y | Y | N | 8 | 14 | 0.57 |
| Congdon 2021 | Y | Y | Y | Y | NA | N | N | N | Y | N | Y | Y | NA | N | NA | NA | 7 | 12 | 0.58 |
| UK National Statistical Bulletin 2021 | Y | Y | Y | N | NA | N | N | N | N | N | Y | Y | NA | N | NR | NR | 5 | 14 | 0.36 |
| Knipe 2021 | Y | Y | Y | Y | NR |  | N | N | Y | N | Y | Y | NA | N | NA | NA | 7 | 13 | 0.54 |
| Sakelliadis 2020 | Y | Y | Y | Y | NR |  | N | N | Y | N | Y | Y | NA | N | NA | NA | 7 | 13 | 0.54 |

N, no; NA, not applicable; NR, not reported; Y, Yes

b) **Case series**

For the list of questions assessed for each type of studies, see <https://jbi-global-wiki.refined.site/space/MANUAL/3290006117/Appendix+7.3+Critical+appraisal+checklists+for+case+series>

|  | **Author Year** | | **Q1** | | **Q2** | | **Q3** | | **Q4** | | **Q5** | | **Q6** | | **Q7** | | **Q8** | | **Q9** | | **Q10** | | **Total score** | | **Total applicable** | | **Ratio** |
| --- | --- | --- | --- | --- | --- | --- | --- | --- | --- | --- | --- | --- | --- | --- | --- | --- | --- | --- | --- | --- | --- | --- | --- | --- | --- | --- | --- |
| Choudhury 2020 | | Y | | Y | | Y | | Y | | Y | | Y | | Y | | NA | | Y | | N | | 8 | | 9 | | 0.89 | |
| Yip 2020 | | Y | | N | | Y | | Y | | Y | | N | | Y | | NA | | Y | | NA | | 6 | | 8 | | 0.75 | |
| Simani 2020 | | Y | | Y | | N | | Y | | Y | | Y | | Y | | NA | | Y | | Y | | 8 | | 9 | | 0.89 | |

N, no; NA, not applicable; NR, not reported; Y, Yes

c) **Case reports**

For the list of questions assessed for each type of studies, see <https://jbi-global-wiki.refined.site/space/MANUAL/3290006119/Appendix+7.4+Critical+appraisal+checklist+for+case+reports>

| **Author Year** | **Q1** | **Q2** | **Q3** | **Q4** | **Q5** | **Q6** | **Q7** | **Q8** | **Total score** | **Total applicable** | **Ratio** |
| --- | --- | --- | --- | --- | --- | --- | --- | --- | --- | --- | --- |
| Forouzanfar 2020 | Y | N | Y | Y | NA | NA | NA | Y | 4 | 5 | 0.80 |
| Pirnia 2020 | Y | N | N | N | NA | NA | NA | Y | 2 | 5 | 0.40 |
| Uğurlu 2020 | Y | Y | Y | Y | NA | NA | NA | Y | 5 | 5 | 1.00 |
| Dumollard 2021 | Y | N | Y | Y | NA | NA | NA | Y | 4 | 5 | 0.80 |

N, no; NA, not applicable; NR, not reported; Y, Yes
